# Surgery versus Balloon angioplasty for treating coarctation of aorta: A meta-analysis

**DOI:** 10.1101/2023.04.06.23288276

**Authors:** Avichal Dani, Helly Thakkar, Dev Desai, Jagdeepkaur S. Dani, Sameer I. Dani

## Abstract

**Background:** In the past 50 years, therapeutic options for treating both children and adults with native aortic coarctation have significantly improved. In contrast to surgery, In 1982, balloon angioplasty (BA) was suggested as a potential alternative for the Primary treatment of CoA. Here, Surgery vs Angioplasty is compared to understand the rates of their compilations like aneurysm and recoarctation.

**Method:** A total of 13 RCTs with a total of 877 patients (Surgery-537, Angioplasty-340) of total patients were identified following PRISMA guidelines till November 2019 and were matched for inclusion and exclusion criteria. The following search strings and MESH terms were used: ‘coarctation of aorta’, ‘surgery’, ‘balloon angioplasty, and ‘aneurysm’. Following this, Surgery and Angioplasty were evaluated for complications and recoarctation. RevMan 5.3 was used for appropriate statistical tests. Fixed and Random Effect Model tests were used and p<0.05 was considered statistically significant.

**Result:** Angioplasty seems to be a Statistically better alternative with lesser complications (OR=1.993, CI95=1.126 to 3.527, p=0.018). It can be seen that Surgery is statistically far better in preventing the formation of an aneurysm (OR=0.291, CI95=0.141 to 0.602, p=0.001). Surgery as a treatment is statistically better than angioplasty to prevent a recoarctation (OR=0.375, CI95=0.268 to 0.524, p=<0.001).

**Conclusion:** Surgery is found to be a better treatment option for preventing complications whereas angioplasty is better in preventing the formation of aneurysms and recoarctation.

## Introduction

Johannes Baptista Morgani initially reported coarctation of the aorta (CoA), a malformation, in 1761. (1) In thee past 50 years, therapeutic options for treating both children and adults with native aortic coarctation have significantly improved. Drs. Crawford and Nylin conducted the initial operation in 1944, and the surgical technique was improved throughout the subsequent four decades. (2)Despite the fact that surgical intervention’s long-term results have been deemed adequate, In 1982, balloon angioplasty (BA) was suggested as a potential alternative to surgery for the Primary treatment of CoA. (1)

In the 1990s, after the transcatheter balloon method, treating this lesion with intravascular stent therapy gained more recognition. Children and adults with native aortic coarctation are treated of choice at several hospitals using the transcatheter method. (2)

The indication for BA has become more clear as experience has grown, with patients who have long-segment coarctation and are younger than three months often being precluded from this kind of intervention. Surgery and BA both experience post-operative recoarctation and aneurysm development. It has not yet been determined if BA should be used as a first-choice treatment over surgical repair in a particular patient population. There are little data contrasting surgical treatment with balloon therapy. Actually, unfortunately, follow-up for all 3 types of treatment has been limited, making it difficult to draw any meaningful conclusions as to which treatment option is superior. (2)

The objective behind this meta-analysis is to find a better treatment modality with less complication for the treatment of Coarctation of the Aorta and to see the outcome results of each of this techniques. The two techniques to be compared here are surgery and angioplasty for rates of different complications with these procedures for example chances of producing an aneurysm, recoarctation, or other complications.

## Methodology

### SEARCH STRATEGY

A search was done using the databases-PubMed, Google Scholar, and Cochrane library for all relevant literature. Full -Text Articles written only in English were considered.

The medical subject headings (MeSH) and keywords ‘coarctation of aorta’, ‘surgery’, ‘balloon angioplasty’, and ‘aneurysm’ were used. References, reviews, and meta-analyses were scanned for additional articles.

### STUDY SELECTION

Titles and abstracts were screened, and Duplicates and citations were removed. References of relevant papers were reviewed for possible additional papers. Papers with detailed patient information and statically supported results were selected.

we searched for papers that complications post procedures for coarctation, where the procedures considered were Surgery or Angioplasty.

The inclusion criteria were as follows: (1) studies that provided information about complications post procedures; (2) studies published in English; (3) Studies comparing Surgery and Angioplasty as a treatment modality for Coarctation

The exclusion criteria were as follows: (1) articles that were not full text ; (2) unpublished articles.

### DATA EXTRACTION

Each qualifying paper was independently evaluated by two reviewers. Each article was analyzed for the number of patients, their age, procedure modality, and incidence of the predecided complications. Further discussion or consultation with the author and a third party was used to resolve conflicts. The study’s quality was assessed using the modified Jadad score. In the end, a total of 13 RCTs with a total of 877 patients were selected according to PRISMA. Out of these 877 patients 537 had Surgery while 340 patients had Angioplasty.

### STATISTICAL ANALYSIS

All of the data was obtained and entered into analytic software. Fixed-or random-effects models were used to estimate mean difference, standardized mean difference (SMD), odds ratios, and relative risk (RR) with 95 percent confidence intervals to examine critical clinical outcomes (CIs). Statistical heterogeneity was measured with the χ2; P < 0.100 was considered as a representation of significant difference. I^2^ greater than or equal to 50% indicated the presence of heterogeneity. Funnel plots were used to assess potential publication bias based on the prevalence of wound infection after surgery. A statistically significant difference was defined as P<0.05.

## RESULT

With the results presented above, it can be seen that when complications are compared between these two modalities of surgery and angioplasty, as per figure 2 Angioplasty seems to be a Statistically better alternative with lesser complications (OR=1.993, CI95=1.126 to 3.527, p=0.018). These results are supported by (3) (4) (5) but (6) says otherwise.

**Figure 1.**
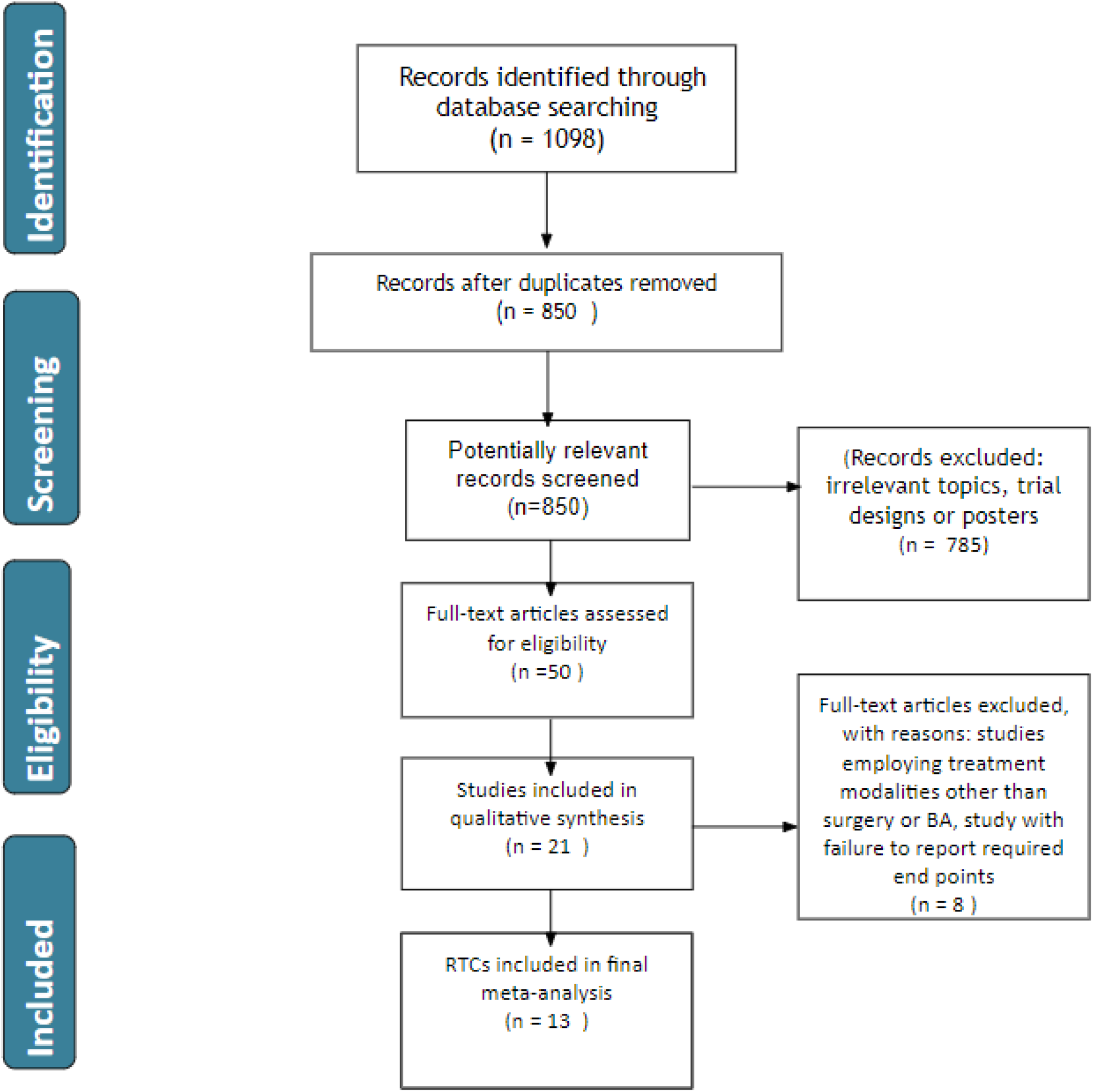
PRISMA Flowchart: -

**Figure 2.**
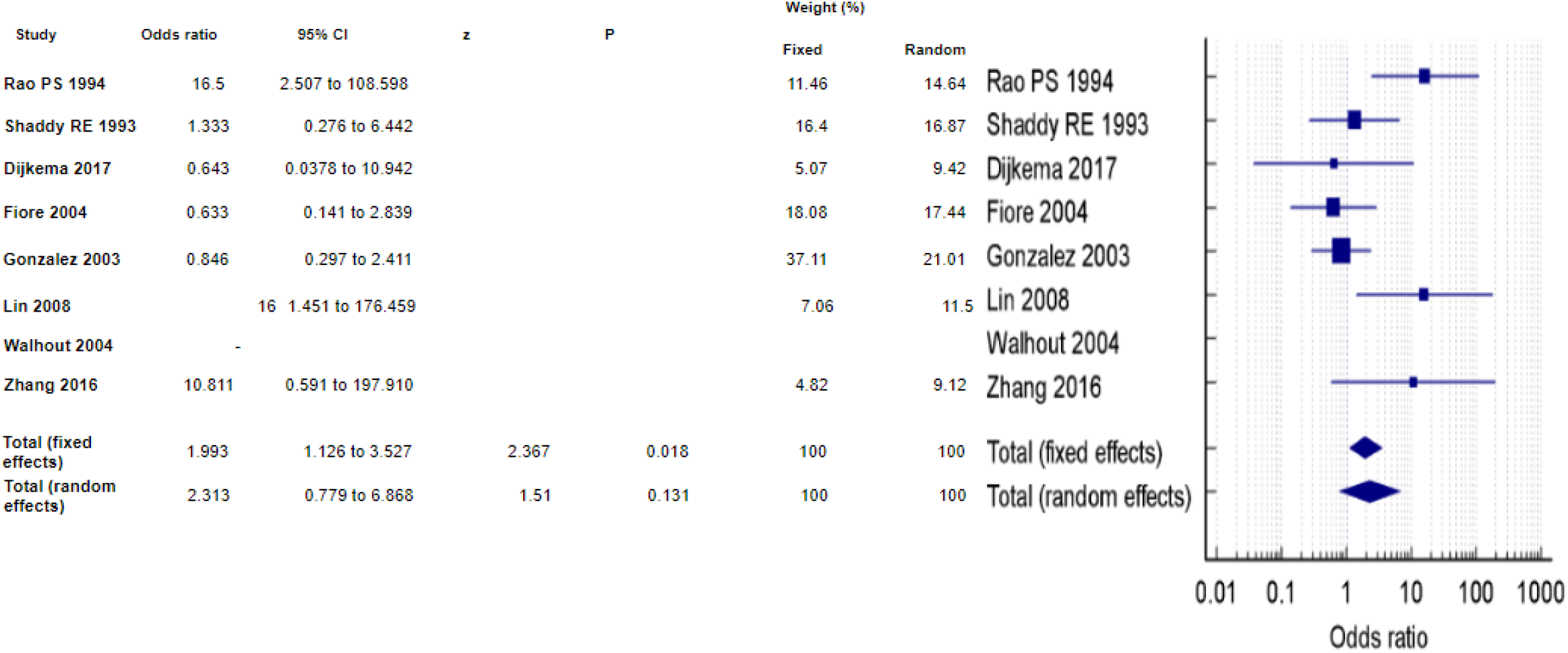
Complications

When the comparison of aneurysms is checked as depicted in figure 3, it can be seen that Surgery is statistically far better in preventing the formation of an aneurysm (OR=0.291, CI95=0.141 to 0.602, p=0.001) and it can be supported by (5) (6) (7) but (8) is the only paper we found saying otherwise and its results are statistically insignificant.

**Figure 3.**
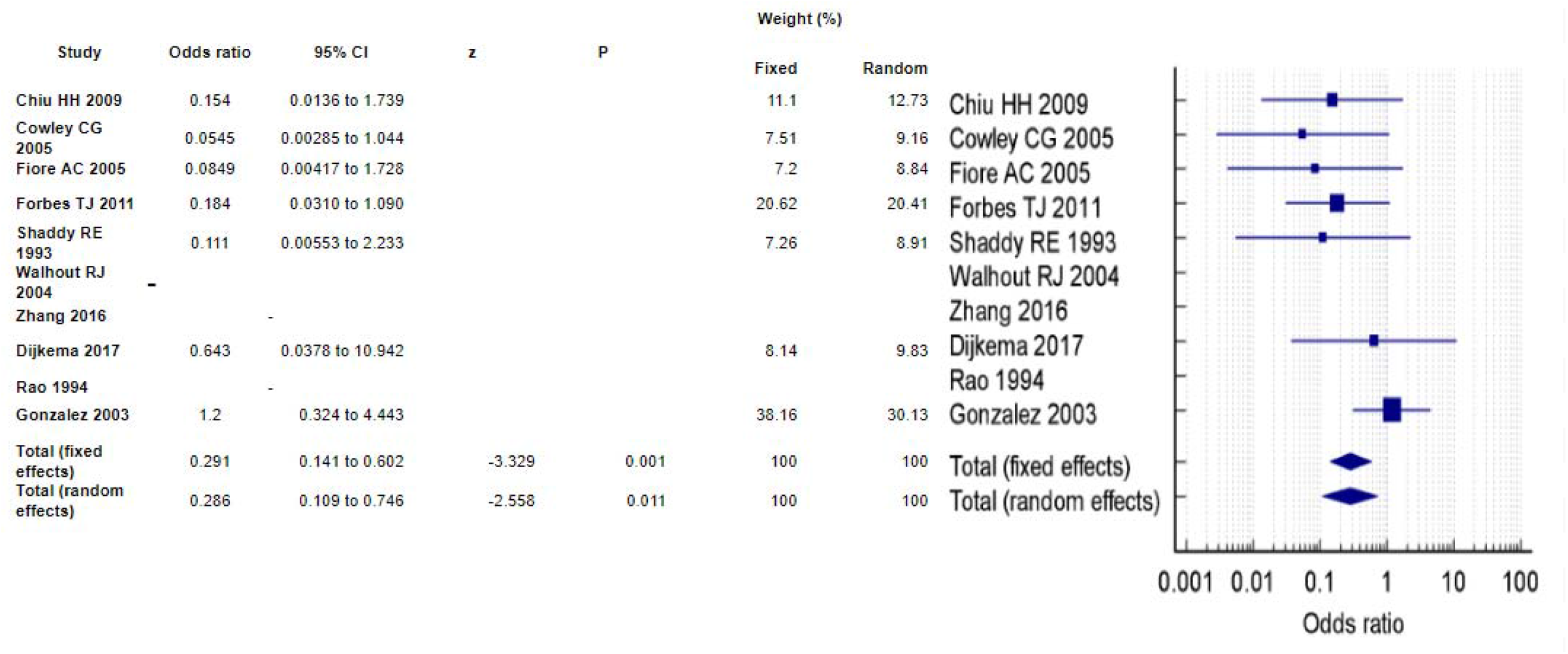
Aneursm

Figure 4 denotes the chances of formation of a coarctation again and it can be seen that surgery as a treatment is statistically better than angioplasty to prevent a recoarctation (OR=0.375, CI95=0.268 to 0.524, p=<0.001). These results are identical to all papers (6) (5) except in (9).

**Figure 4.**
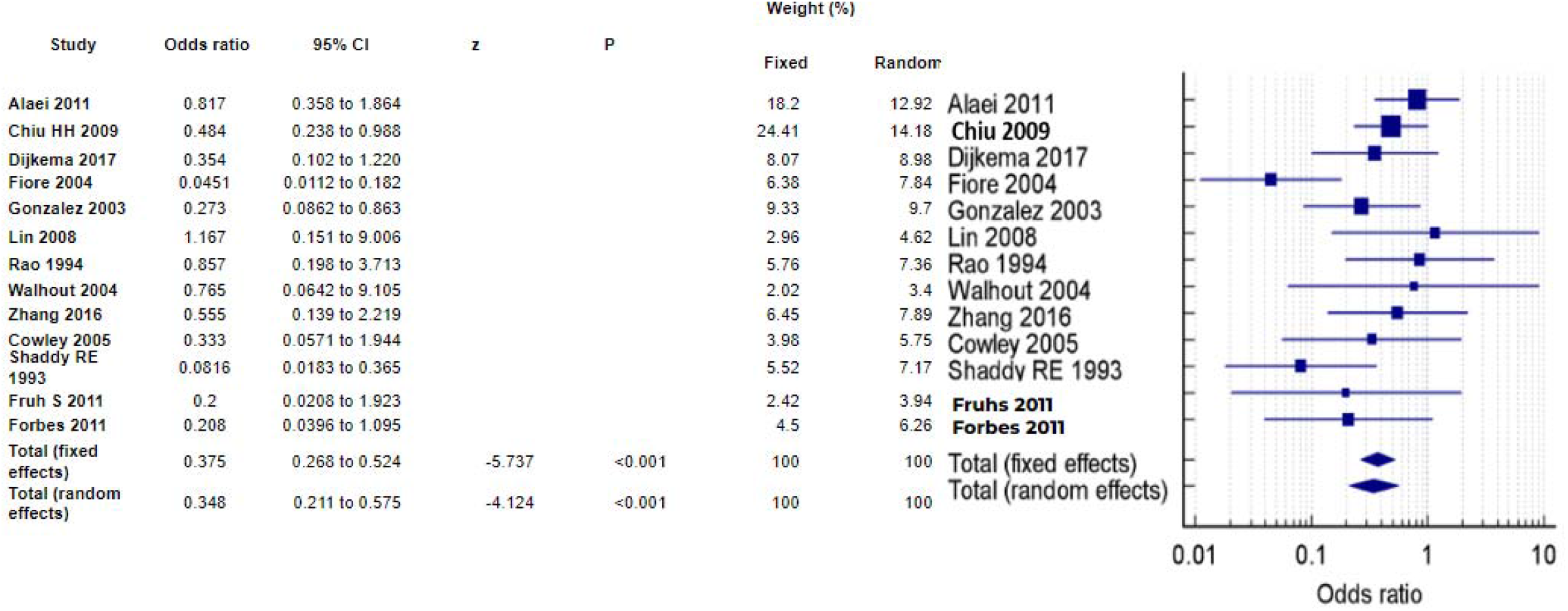
Re-CoA

## Discussion

This seems like a common pattern in almost all literature present anywhere is that out of all the modalities present to treat a coarctation, be it Surgery or Angioplasty with balloon or stent; no modality is full proof and each procedure has its own fallings when it comes to the complications it produces in the patients.

It is seen in almost all patients that surgery as a treatment modality beings the highest amount of post-operation complications which include (3) (4). In contrast to Surgery, we found that angioplasty produces very low incidences of these complications and these results are similar to many research papers present in the current literature available. There are some papers present that shows that angioplasty might carry higher chances of these complications but it can be seen that most of those papers have statistically insignificant results.

An aneurysm is one of the major complications produced in a treated coarctation as the wall is damaged and weak. The outpouching of this weak wall produces an aneurysm which can rupture and cause a big hemorrhage and hematoma. Failure to manage this patient almost results in the death of the patient. Aneurysm and rupture of these aneurysms in the brain can lead to stroke and sudden death. It is evident with the presented results that Angioplasty actually decreases the odds of producing an aneurysm in treated coarctation in contrast to surgery(6, 7); maybe because surgery opens the vessel wall so when stitched back, it is ought to bring some kind of weakness due to that not being the original tissue but being a healed tissue. Angioplasty is repeatedly found to be a better treatment modality in preventing an aneurysm formation.

One of the major issues with treating a coarctation is that the treated region might form a coarctation again. Also known as recoarctation, it is a major complication and needs the usage of treatment again. It was found in our results that angioplasty is far superior in preventing the formation of a recoarctaion and this is similar to the pattern found in present literature and studies(5, 6). The reason behind a higher rate of recoarctation with surgery is that when the vessel is closed again, there is bound to become some type of stricture as the normally elastic tissue will be replaced with healing tissue.

### Conclusion

All treatment modalities for the treatment of Coarctation have their own issues when it comes to complications but Surgery is found to be a better treatment option for preventing complications whereas angioplasty is better in preventing the formation of aneurysm and recoarctation.

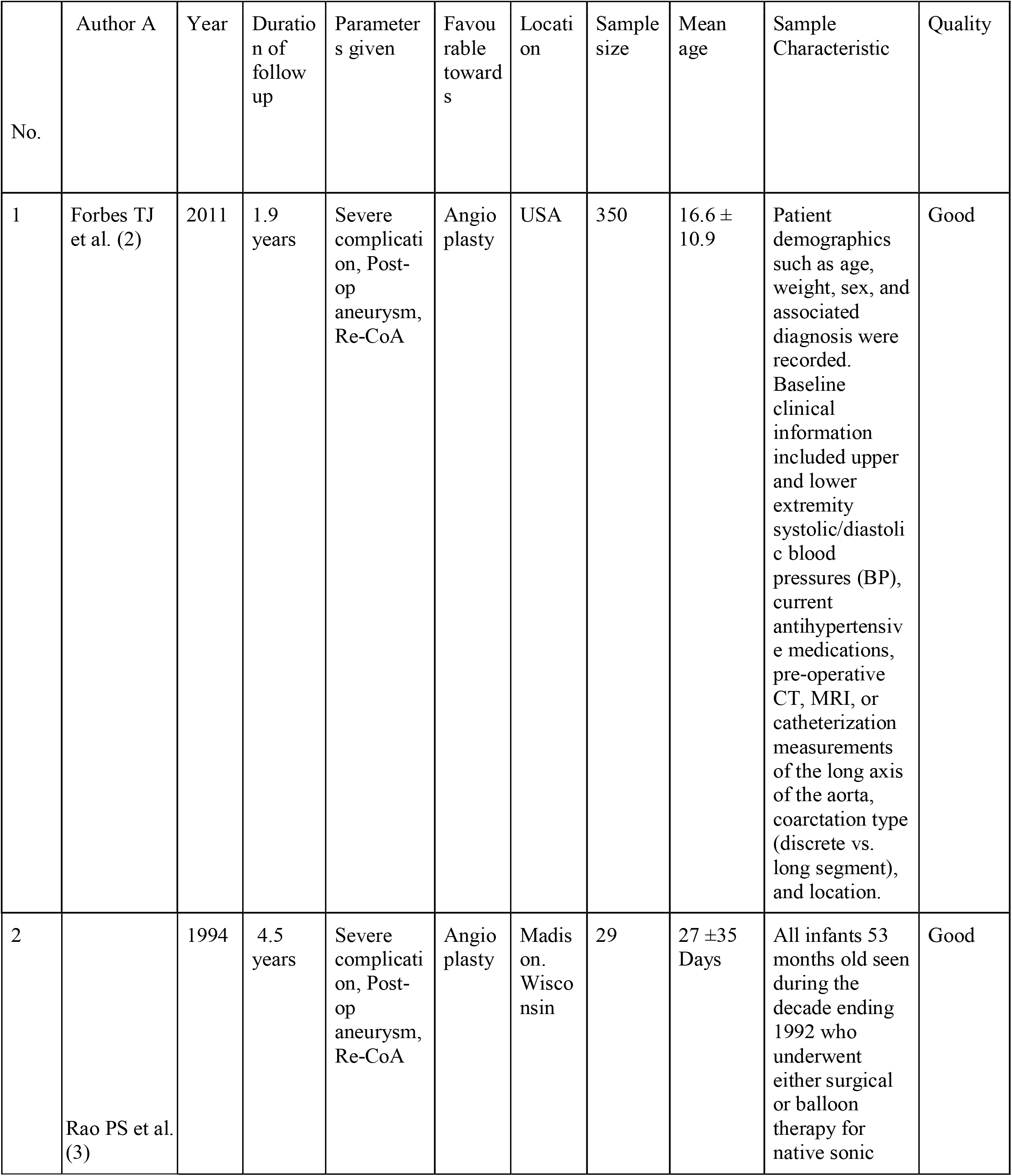
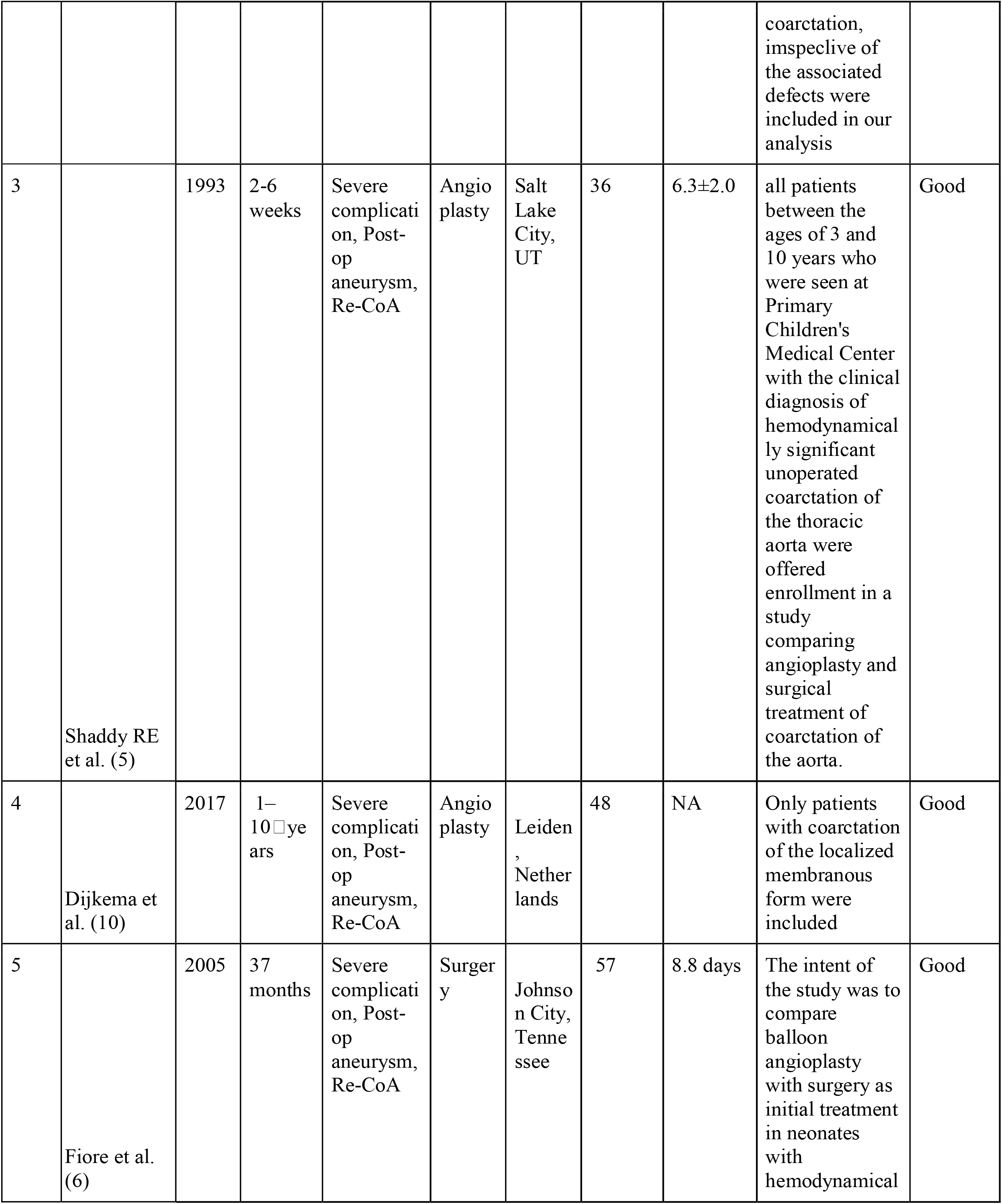
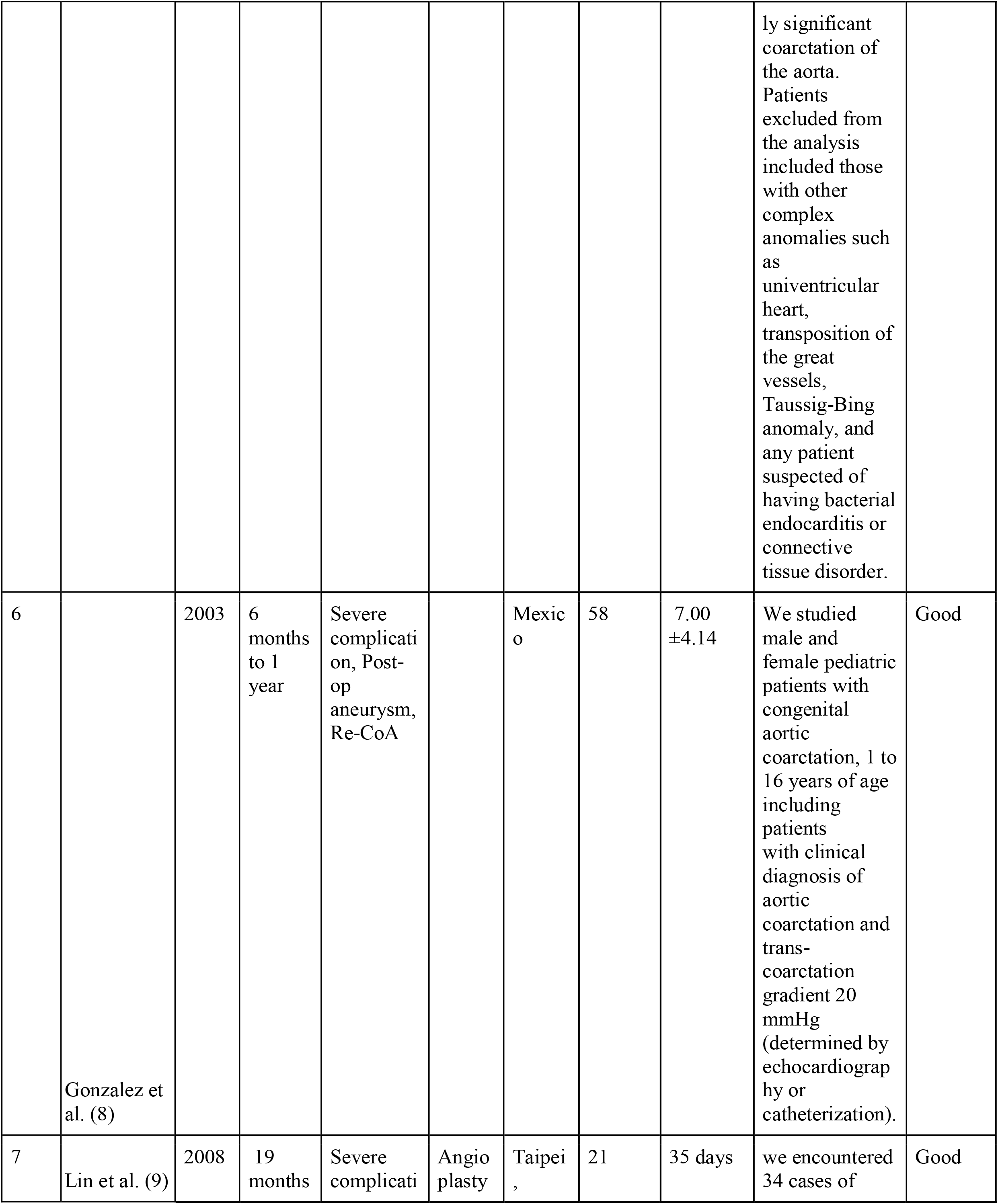
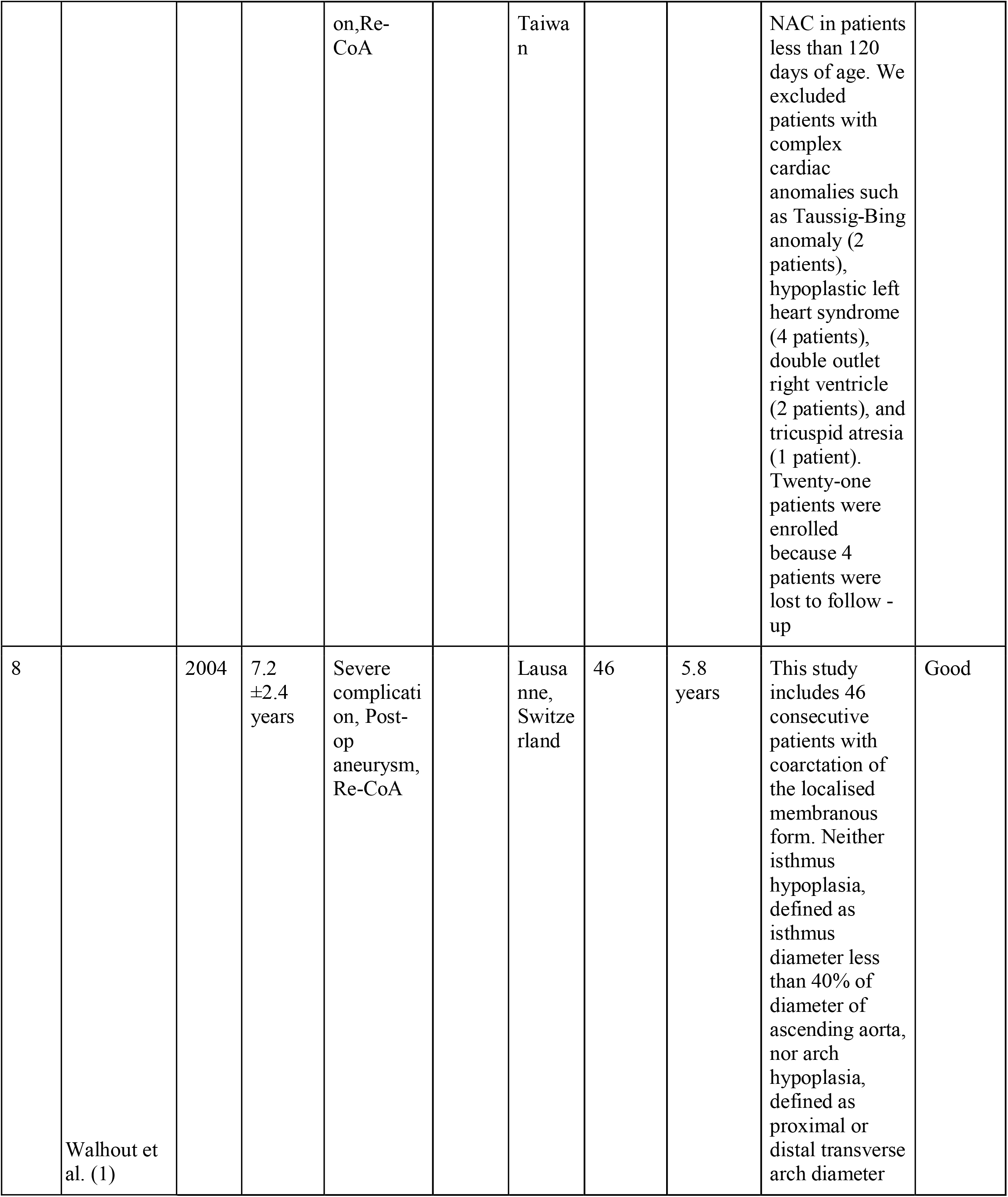
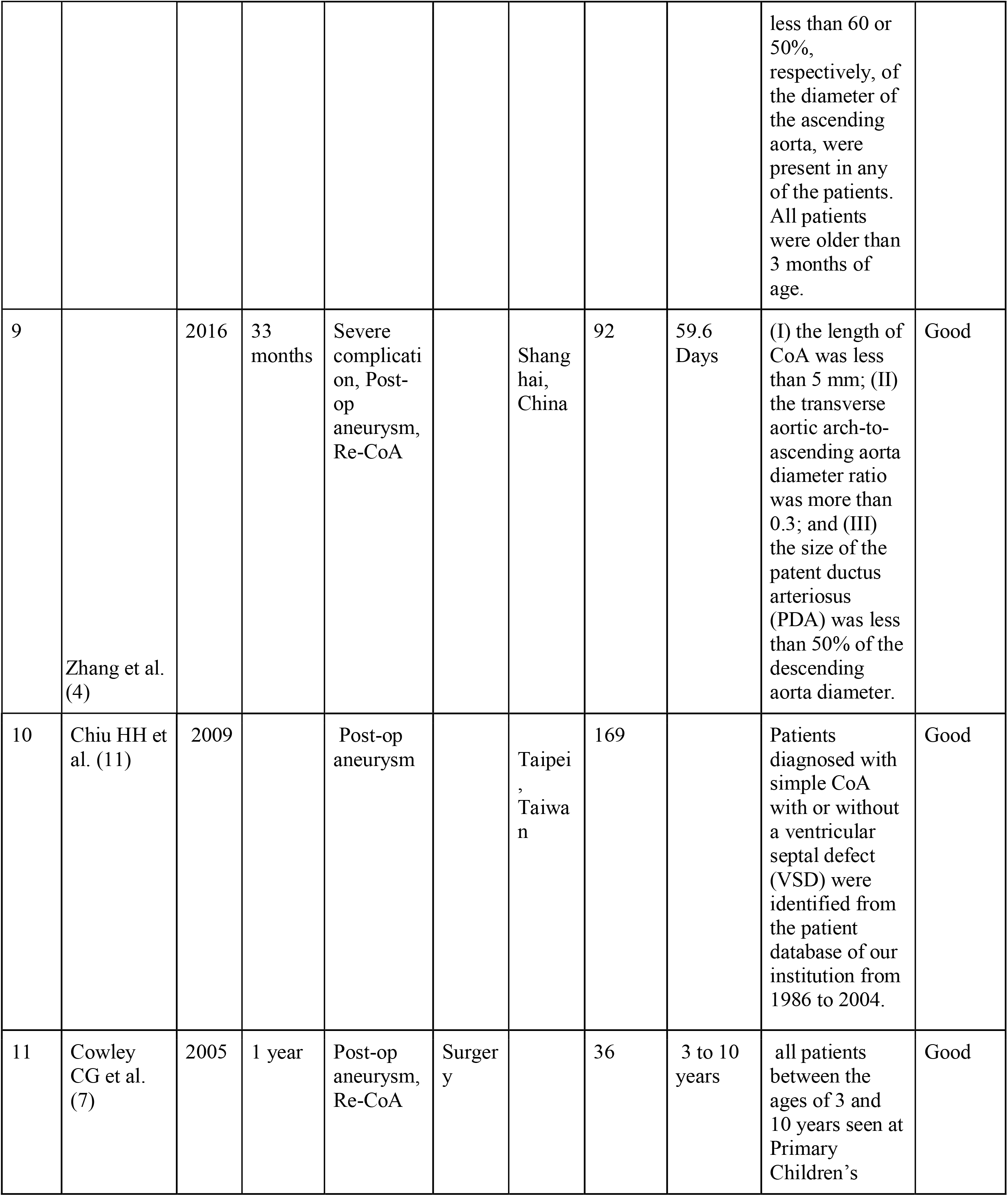
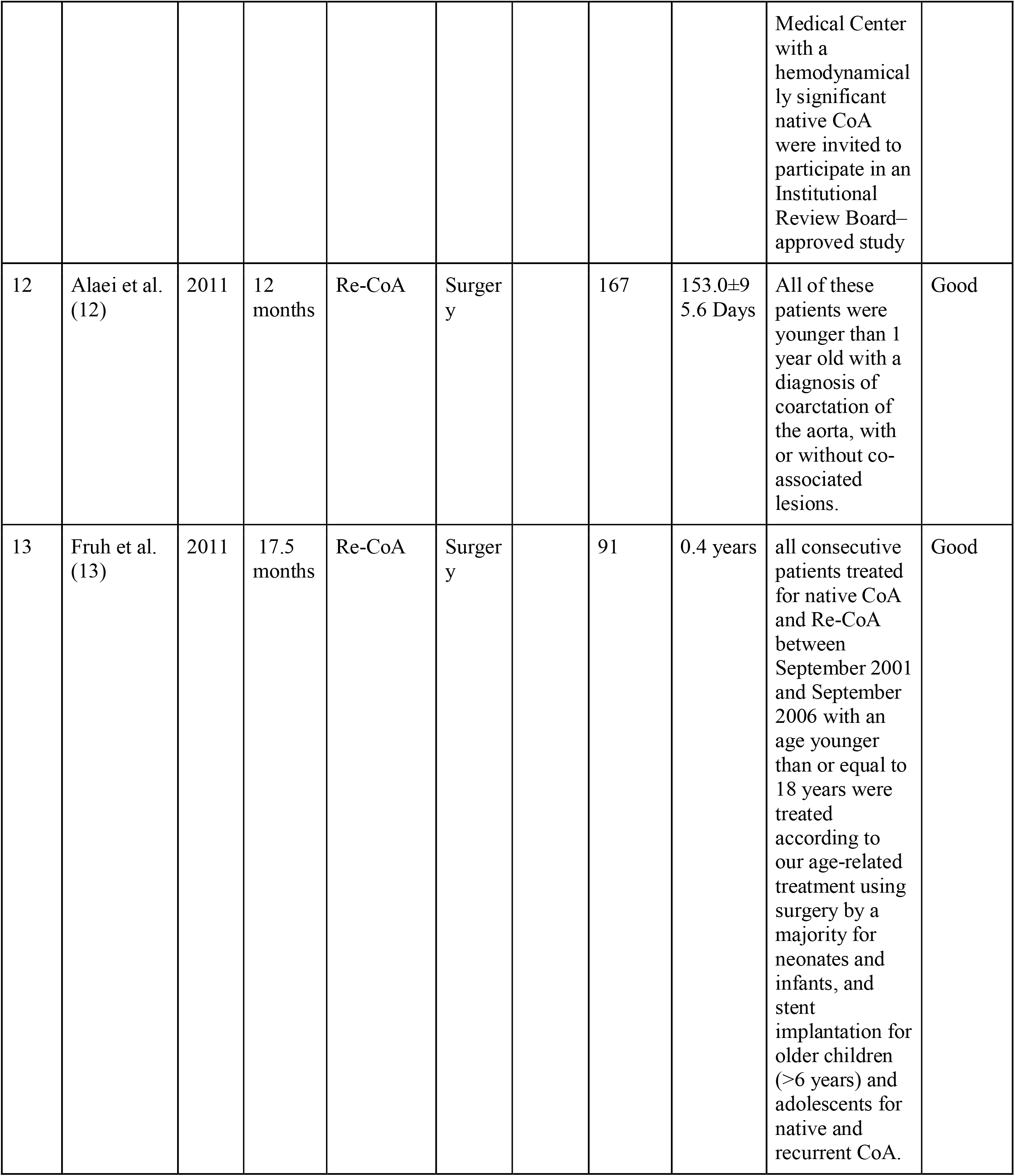

## Data Availability

All data produced in the present work are contained in the manuscript

---

**Supplemental Table 2**

