## Supplemental Table 2 for "Surgery versus Balloon angioplasty for treating coarctation of aorta: A meta-analysis"

### **Supplementary table.2** Summary of postoperative complications, aneurysm formation and perioperative mortality between BA and surgery

| Study | Surgical approach | Complications |
| --- | --- | --- |
| Shaddy RE(1) | SU  BA | 2 diminished pulse, 1 bleeding, 1 hypertension  2 bleeding, 1 paraparesis, 1 vocal cord paralysis |
| Zhang(2) | SU  BA | 6 recurrent laryngeal nerve injury  no complication observed |
| Lin(3) | SU  BA | 1 chylothorax; 1 sepsis;1 tracheal stenosis; 3 atelectasis;1 wound infection;1 pneumonia  1 intimal tear of the aorta |
| Dijkema(4) | SU  BA | 1 bleeding  1 thrombosis |
| Walhout(5) | SU  BA | NA  NA |
| Fiore(6) | SU  BA | 1 bleeding; 3 chylothorax  1 tamponade; 3 thrombosis |
| Gonzalez(7) | SU  BA | 2 bleeding; 1 hypertension crisis; 1 diaphragmatic nerve injury; 3 pulmonary sepsis;  1 cardiogenic shock; 2 dehiscence sutures; 1 chylothorax  3 bleeding; 1 hypertension crisis;3 thrombosis; 1 anaphylatic shock; 1 cardiogenic shock;  4 inguinal hematoma |
| Rao(8) | SU  BA | 1 seizure;1 right middle lobe infarction; 2 cardiac arrest; 2 pneumothrax; 2 septicemia  2 femoral artery injury; 2 bleeding |

*SU* surgery, *BA* balloon angioplasty group, *NA* not available

References:-
